# Noncommunicable diseases associated with hemorrhagic disorders, hospitalization and mortality in patients with dengue in Mexico: a national analysis of confirmed cases in 2024

**DOI:** 10.1101/2025.02.24.25322761

**Authors:** Diego Rolando Hernández Galdamez, Miguel Ángel González-Block, Daniela Karola Romo-Dueñas, Erick Antonio Osorio López, Rosalba Cerón-Meza, Pablo Méndez-Hernández

## Abstract

**Background:** In Mexico, some of the most prevalent non-communicable diseases (NCDs) among adults are diabetes, hypertension, and chronic kidney disease (CKD). Mexico could be going through a syndemic of dengue and NCDs. This study aims to describe and analyze the association between the prevalence of NCDs and hospitalization, the presence of hemorrhagic disorders, and death in all officially confirmed cases of dengue in Mexico during 2024.

**Methodology/Principal Findings:** This cross-sectional study is carried out through a secondary analysis of the confirmed cases of dengue reported in 2024. The likelihood of hospitalization, hemorrhagic disorders, and death were assessed according to NCDs, estimating odds ratios (ORs). A multivariate logistic regression model was used, adjusting by age, sex, and for each of the NCDs (diabetes, hypertension, chronic kidney disease, immunosuppression, cirrhosis, and peptic ulcer disease). The most co-prevalent non-communicable diseases were diabetes, hypertension, CKD, and immunosuppression. For hospitalization, CKD has the highest risk of hospitalization (OR 5.73), followed by immunosuppression (OR 2.84), peptic ulcer disease (OR 2.35), and diabetes (OR 2.09). We found higher probabilities for having bleeding disorders in people with diabetes (OR 19.51), peptic ulcer disease (OR 7.85), immunosuppression (OR 4.75), cirrhosis (OR 3.37), and hypertension (OR 2.99) compared to people who did not have these diseases. The highest risk for death was observed in the cases with CKD (OR 6.54), followed by peptic ulcer disease (OR 3.09) and diabetes (OR 2.87).

**Conclusions/Significance:** Patients with dengue and some comorbidities were more likely to have worse clinical outcomes, such as being hospitalized, having bleeding disorders, or dying. The syndemic of NCDs and dengue in Mexico has been rapidly increasing, and this problem needs to be addressed. This work demonstrates that patients with these comorbidities have worse clinical outcomes.

## Introduction

Dengue is a viral infection caused by Dengue flavivirus (DENV), which belongs to the RNA Flaviviridae virus family. DENV commonly hosts mosquitoes as vectors of these species, like Aedes aegypti, transmit the DENV to humans (1). These mosquitoes are spreading their reach due to climate change and global warming, posing disease control challenges in areas previously free from dengue. (2). Dengue is considered a neglected tropical infection with high endemicity, especially in Latin America (3). According to the Pan American Health Organization (PAHO), 2024 is outlined to record the highest number of dengue cases since 1980. For Latin America, this represents a 70% increase in reported confirmed cases from 2023 (2,053,822) to 2024 (6,916,684). Mexico reported a rise of 126.3% in confirmed cases from 2023 (54,406) to 2024 (123,142) (4). Brazil has the highest incidence in the Americas (5).

Severe dengue (SD) is a complication that generally occurs when the fever goes away and is accompanied by intense abdominal pain, vomiting, bleeding in the gums or nose, or vomiting blood or blood in the stool, among other symptoms. These are known as warning signs (WS) that require immediate medical attention, which may or may not require hospitalization (6). Previous studies have identified some characteristics that can serve as predictors for developing severe dengue. A 2021 meta-analysis that included studies from the Americas identified children, secondary infection, diabetes, and renal disease as predictors for severe dengue (7). Another study considers chronic kidney disease (CKD) as a high-risk factor for developing WS and SD, especially in those with end-stage renal disease on maintenance dialysis (8).

Mexico could be going through a syndemic of neglected diseases, such as dengue and non-communicable diseases (NCD). In the Mexican context, the prevalence of NCDs among adults is diabetes (18.3%), hypertension (29.9%), and CKD (9.11%) (9–11). There is solid evidence that the presence of pre-existing NCD triggers severe illness and increases the risk of mortality in those who get severe dengue (12,13). A recent study in México identified that diabetes and CKD were associated with increased mortality risk (14).

Identifying predictive factors of severe dengue disease is useful in prioritizing interventions for preventing dengue transmission and urgent measures for NCD prevention and control (13). This study aims to describe and analyze the association between the prevalence of NCDs and hospitalization, the presence of hemorrhagic disorders, and death in all official confirmed dengue cases reported in Mexico during 2024.

## Methods

### Databases and data extraction

This cross-sectional study is carried out through a secondary analysis of the confirmed dengue cases reported by the Federal Ministry of Health (MoH) of Mexico through the anonymized and open-access database published through the Directorate General of Epidemiology.

The MoH database, from January 1, 2024, to December 4, 2024, reported 117,495 laboratory-confirmed dengue cases. The following variables were extracted and assessed. For sociodemographic status: age, sex, ethnicity, and social security affiliation. For noncommunicable diseases: diabetes, hypertension, chronic kidney disease, immunosuppression, cirrhosis, and peptic ulcer disease. For outcome variables: hospitalization, presence of hemorrhagic disorders, and deceased. The descriptors in the database do not define the classification method for comorbidities. The information is obtained through a dichotomous questionnaire that the interviewer fills with the information provided by the patient.

The study does not require ethical review because it is based on open, anonymized data from the Mexican Ministry of Health.

### Statistical Analysis

Categorical variables were described as percentages. χ2 and Fisher’s exact tests were performed to compare the proportions of patients with and without noncommunicable diseases against the proportions of patients with and without hospitalization, hemorrhagic disorders, and death. The likelihood of hospitalization, hemorrhagic disorders, and death were assessed according to pre-existing medical conditions, estimating odds ratios (ORs) with 95% confidence intervals and their corresponding p values. A multivariate logistic regression model was used, adjusting by age, sex, and for each of the noncommunicable diseases (diabetes, hypertension, chronic kidney disease, immunosuppression, cirrhosis, and peptic ulcer disease). All statistical analysis was performed using Stata MP software, version 14.0 (Stata Corporation, College Station, TX, USA).

## Results

Of the total confirmed dengue cases in 2024, 55.66% were women. The age group with the highest concentration of cases was adults aged 20 to 44 (41.83%), followed by adolescents (26.52%). Only 2.42% of reported cases occurred in Indigenous people. 46.04% of the population have social security (IMSS, ISSSTE, Sedena, Pemex, or Semar). The most co-prevalent non-communicable diseases were diabetes (2.97%), hypertension (2.47%), CKD (0.26%), and immunosuppression (0.17%) (Table 1).

**Table 1.** General characteristics of patients with dengue and condition of hospitalization, hemorrhagic disorders, and mortality, Mexico 2024.

| Variables | Total confirmed cases<br>n=117,495 | In-patient cases<br>n= 45,568 |  | Cases with hemorrhagic disorders<br>n=1,028 |  | Deceased cases<br>n= 776 |  |
| --- | --- | --- | --- | --- | --- | --- | --- |
|  | n (%) | n (%) | p | n (%) | p | n (%) | p |
| **Sex** |  |  |  |  |  |  |  |
| Women | 65,401(55.66) | 25,913 (56.87) | 0.000* | 630 (61.28) | 0.000* | 428 (55.15) | 0.775* |
| Men | 52,094 (44.34) | 19,655 (43.13) |  | 398 (38.72) |  | 348(44.85) |  |
| **Age group** |  |  |  |  |  |  |  |
| < 5 | 3,077 (2.62) | 1,439 (3.16) | 0.000* | 4 (0.39) | 0.000* | 31 (3.99) | 0.000* |
| 5-9 | 9,324 (7.94) | 4,158 (9.12) |  | 11 (1.07) |  | 52 (6.70) |  |
| 10-19 | 31,161 (26.52) | 13,987 (30.69) |  | 32 (3.11) |  | 119 (15.34) |  |
| 20-44 | 49,154 (41.83) | 16,804 (36.88) |  | 245 (23.83) |  | 242 (31.19) |  |
| 45-59 | 16,186 (13.78) | 5,276 (11.58) |  | 395 (38.42) |  | 112 (14.43) |  |
| >60 | 8,593 (7.31) | 3,904 (8.57) |  | 341 (33.17) |  | 220 (28.35) |  |
| **Ethnicity** |  |  |  |  |  |  |  |
| Indigenous | 2,843 (2.42) | 860 (1.89) | 0.000* | 32 (3.11) | 0.146* | 21(2.71) | 0.558** |
| No Indigenous | 114,652 (97.58) | 44,708 (98.11) |  | 996 (96.89) |  | 755 (97.29) |  |
| **Social security affiliation** |  |  |  |  |  |  |  |
| With social security | 54,089 (46.04) | 20,394 (44.76) | 0.000* | 548 (53.31) | 0.000* | 283 (36.47) | 0.000* |
| Without social security | 63,406 (53.96) | 25,174 (55.24) |  | 480 (46.69) |  | 493 (63.53) |  |
| **Comorbidity** |  |  |  |  |  |  |  |
| Diabetes | 3,488 (2.97) | 1,930 (4.24) | 0.000* | 461 (44.84) | 0.000* | 123 (15.85) | 0.000* |
| Hypertension | 2,906 (2.47) | 1,527 (3.35) | 0.000* | 22 (2.14) | 0.490* | 81 (10.44) | 0.000** |
| Chronic kidney disease | 310 (0.26) | 253 (0.56) | 0.000* | 24 (2.33) | 0.000** | 33 (4.25) | 0.000** |
| Immunosuppression | 196 (0.17) | 131 (0.29) | 0.000* | 13 (1.26) | 0.000** | 6 (0.77) | 0.002** |
| Cirrhosis | 95 (0.08) | 55 (0.12) | 0.000* | 6 (0.58) | 0.000** | 2 (0.26) | 0.131** |
| Peptic ulcer disease | 59 (0.05) | 40 (0.09) | 0.000* | 11 (1.07) | 0.000** | 5 (0.64) | 0.000** |

\* $\chi^2$ test \*\*Fisher's exact test

According to the data analysis, 38.7% (45,568) of the confirmed dengue cases were hospitalized, among which hospitalized women were 56.87%. In the case of hospitalized patients, the most frequent age group was 20-44 years (36.88%), followed by 10-19 years (30.69%). 55.24% of the in-patients’ cases didn’t have social security affiliation. We found a higher prevalence of NCDs in hospitalized patients compared to outpatients. We found a higher prevalence of NCDs in hospitalized patients compared to outpatients: diabetes (4.24% vs. 2.17%), hypertension (3.35% vs 1.92%), and CKD (0.56% vs 0.08%). It was reported that 0.87% (1,028) of confirmed cases presented hemorrhagic disorders, of which 61.28% were women. In this case, the age groups with the highest incidence of bleeding disorders were adults aged 45 to 59 (38.42%) and adults over 60 (33.17%). Among those who had hemorrhagic disorders, 44.84% had diabetes, and 2.88% had CKD as the most prevalent comorbidity. No significant differences were found concerning sex or ethnicity. Regarding mortality, 63.53% of them didn’t have social security affiliation. The age group in which most fatal cases occurred was young adults aged 20 to 44 (31.19%), followed by older adults (28.35%). Among patients with diabetes and hypertension, we observed higher mortality rates compared to the population without these NCDs (3.53% vs 0.57% and 2.79% vs 0.61%, respectively). The most prevalent comorbidities in patients who died were diabetes (15.85%), hypertension (10.44%), and CKD (4.25%) (Table 1).

Regarding sociodemographic variables, we found significant differences in mortality rates between people with social security and people who did not have social security (36.5 vs 63.5%, respectively). On the other hand, the indigenous population had a lower hospitalization rate in comparison to no indigenous people (30.2% vs 38.9%), despite the similarity in mortality rates between the groups.

Patients with dengue and some comorbidities were more likely to have worse clinical outcomes, such as being hospitalized, having bleeding disorders, or dying. For hospitalization, the analysis reported that women were 1.10 times more likely to be hospitalized than men. A higher risk was reported for patients older than 60 (OR 1.19) compared with children under 5. The rest of the age groups between 5 and 59 years old were less likely to be hospitalized than children under 5 (ORs from 0.97 to 0.65). Among NCDs, CKD has the highest risk of hospitalization (OR 5.73), followed by immunosuppression (OR 2.84), peptic ulcer disease (OR 2.35), and diabetes (OR 2.09) (Table 2).

**Table 2.** Adjusted odd ratios for hospitalization, hemorrhagic disorders, and mortality in dengue-confirmed cases in Mexico 2024.

| Variables | In-patient cases<br>n= 45,568 |  |  | Cases with hemorrhagic disorders<br>n=1,028 |  |  | Deceased cases<br>n= 776 |  |  |
| --- | --- | --- | --- | --- | --- | --- | --- | --- | --- |
|  | Adjusted odds ratio | 95% CI | p | Adjusted odds ratio | 95% CI | p | Adjusted odds ratio | 95% CI | p |
| **Sex** |  |  |  |  |  |  |  |  |  |
| Men | 1 |  |  | 1 |  |  | 1 |  |  |
| Women | 1.10 | 1.07-1.13 | 0.000 | 1.19 | 1.05-1.35 | 0.007 | 0.92 | 0.80-1.06 | 0.291 |
| **Age group** |  |  |  |  |  |  |  |  |  |
| <5 | 1 |  |  | 1 |  |  | 1 |  |  |
| 5-9 | 0.93 | 0.86-1.01 | 0.096 | 0.75 | 0.24-2.37 | 0.629 | 0.44 | 0.28-0.68 | 0.000 |
| 10-19 | 0.97 | 0.89-1.04 | 0.406 | 0.51 | 0.18-1.45 | 0.208 | 0.22 | 0.14-0.33 | 0.000 |
| 20-44 | 0.65 | 0.58-0.71 | 0.000 | 1.34 | 0.48-3.75 | 0.577 | 0.13 | 0.07-0.21 | 0.000 |
| 45-59 | 0.65 | 0.57-0.74 | 0.000 | 3.27 | 1.09-9.76 | 0.033 | 0.07 | 0.03-0.14 | 0.000 |
| >60 | 1.19 | 1.01-1.41 | 0.043 | 2.84 | 0.87-9.26 | 0.084 | 0.12 | 0.05-0.27 | 0.000 |
| **Ethnicity** |  |  |  |  |  |  |  |  |  |
| No Indigenous | 1 |  |  | 1 |  |  | 1 |  |  |
| Indigenous | 0.67 | 0.63-0.74 | 0.000 | 1.25 | 0.87-1.79 | 0.226 | 1.11 | 0.72-1.71 | 0.647 |
| **Social security affiliation** |  |  |  |  |  |  |  |  |  |
| With social security | 1 |  |  | 1 |  |  | 1 |  |  |
| Without social security | 1.06 | 1.04- 1.09 | 0.000 | 0.85 | 0.75-0.96 | 0.010 | 1.66 | 1.43-1.92 | 0.000 |
| **Comorbidity\*** |  |  |  |  |  |  |  |  |  |
| Diabetes | 2.09 | 1.94-2.25 | 0.000 | 19.51 | 16.90- 22.53 | 0.000 | 2.86 | 2.26-3.65 | 0.000 |
| Hypertension | 1.59 | 1.47-1.73 | 0.000 | 0.04 | 0.02-0.06 | 0.000 | 1.17 | 0.87-1.56 | 0.296 |
| Chronic kidney disease | 5.73 | 4.28-7.69 | 0.000 | 2.99 | 1.74-5.15 | 0.000 | 6.54 | 4.32-9.89 | 0.000 |
| Immunosuppression | 2.84 | 2.09-3.86 | 0.000 | 4.75 | 2.41-9.33 | 0.000 | 1.76 | 0.14-4.46 | 0.228 |
| Cirrhosis | 1.89 | 1.25-2.88 | 0.003 | 3.37 | 1.16-9.75 | 0.025 | 0.79 | 0.14-4.46 | 0.789 |
| Peptic ulcer disease | 2.35 | 1.32-4.17 | 0.004 | 7.85 | 3.3-18.9 | 0.000 | 3.09 | 1.08-8.88 | 0.036 |

\*Compare without each comorbidity
Odds Ratio were adjusted by age, sex, and for each of the comorbidities analyzed.

Concerning hemorrhagic disorders, women were more likely to have them than men (OR 1.19). The age groups at greatest risk of hemorrhagic dengue were adults aged 45 to 59, followed by older adults (OR 3.27), compared to children under 5 years of age.

Interestingly, when analyzing people with dengue and comorbidities, very high probabilities were found for having bleeding disorders in people with diabetes (OR 19.51), peptic ulcer disease (OR 7.85), immunosuppression (OR 4.75), cirrhosis (OR 3.37) and hypertension (OR 2.99) (Table 2).

Regarding mortality, of the total number of confirmed cases of dengue, 776 people died (0.66%). People without social security had a higher risk of dying (OR 1.66). All the older age groups were less likely to die than children under 5 years of age (ORs from 0.44 to 0.07). The highest risk for death was observed in the cases with CKD (OR 6.54), followed by peptic ulcer disease (OR 3.09) and diabetes (OR 2.87). Finally, we identified those who presented with hemorrhagic disorders had a higher risk of hospitalization (OR 2.67, 95% CI 2.35-3.02) and death (OR 3.70, 95% CI 2.68-5.12). (Table 2)

## Discussion

In this study, we have found consistent findings concerning previous studies. Our results suggest that the presence of pre-existing NCDs in people with dengue is strongly associated with hospitalization, the development of hemorrhagic disorders, or death.

In the case of hospitalization, although they were not the most prevalent, the NCDs that were most associated were CKD, immunosuppression, and peptic ulcer disease; in the case of the presence of bleeding disorders, diabetes, peptic ulcer disease, and immunosuppression; finally, in the case of death, CKD, diabetes and peptic ulcer disease. We also identified women and the extremes of life, i.e., children under five and adults over 60, as a risk group associated with a higher probability of hospitalization. However, adults aged 45 to 59 and over 60 had the highest probability of presenting hemorrhagic disorders, which can be explained by the prevalence of NCDs in this age group compared to children. Our analysis has limitations concerning the variable of “hemorrhagic disorders” given that there is no further information on what was considered to classify the confirmed cases with the presence of these. The lack of information prevents us from classifying the cases as dengue with warning signs or severe dengue, according to Mexican Standards. (15). In the latter document, severe dengue is only considered when there is already shock due to extravasation, severe bleeding (for example, hematemesis, melena), or organ failure. This type of information is not included in the database, so we consider the presence of this condition as a warning sign (WS). Nor can we know if the appearance of these warning signs was before or after hospitalization. However, we consider this variable to be a key predictor and strong proxy for severe dengue because of its association with hospitalization and death, and its association with some NCDs is the main contribution of our study.

Our results are consistent with the findings of other studies identifying CKD and diabetes as the main NCDs for the risk of severe dengue and death (7,8,14). In the case of the study by Rios-Bracamontes et al., our ORs are slightly higher for CKD (6.54 vs. 3.35) and diabetes (2.86 vs. 2.07) (14). It is important how other NCDs, such as peptic ulcer disease, play an important role as they are associated with a higher probability of hospitalization, bleeding, and death. This can be explained by the risk per se that these pathologies have, like cirrhosis, of causing upper gastrointestinal bleeding, and this generates a synergistic factor with dengue (16). Therefore, the antecedent of peptic ulcer disease should be urgently considered by clinicians as a predictor for these outcomes.

Although Mexico has a low case fatality rate (0.66%) compared to other countries in the region, if we compare it with Brazil, which has the highest incidence of cases in the Americas, the latter’s case fatality rate has ranged between 0.04 and 0.09% in the last 10 years (2,17). This reinforces the need for epidemiological surveillance systems for suspicious cases and to classify them according to risk factors to avoid complications. One of the most important early predictors must be the appearance of WS, among the most important being signs of bleeding, according to our study, the NCDs most associated with these hemorrhagic disorders are diabetes, peptic ulcer disease, and immunosuppression, in addition to CKD. To this, we would add being a woman and being in the over-45 age group to identify a risk group that is slightly different from the one usually considered (under 5 years of age). Therefore, the clinical approach following confirmation of a case of dengue should be risk classification according to these predictors and consider early observation/hospitalization.

The National Health Program 2020–2024 proposed intersectoral actions to mitigate the impact of climate change on health, although no specific mention is made of the syndemic between neglected diseases and diseases associated with global warming and NCDs (18). Other countries have proposed specific actions to monitor the impact of global warming among people living with NCDs and to develop health system adaptations to address the syndemic. (19,20).

## Conclusions

The syndemic of NCDs and dengue in Mexico has been rapidly increasing, and this problem needs to be addressed. This work demonstrates that patients with these comorbidities have worse clinical outcomes. Further research is needed to better understand the effect of sociodemographic conditions on clinical outcomes in patients with dengue in Mexico.

Finally, the monitoring of health risks posed by global warming should consider the population living with NCDs, given the higher risk posed by dengue to these populations, as well as the high prevalence of NCDs in Mexico.

## Data Availability

All relevant data are within the manuscript and its Supporting Information files.

https://www.gob.mx/cms/uploads/attachment/file/965318/Datos_abiertos_historicos_etv_2024.pdf

## References

1. Pourzangiabadi M, Najafi H, Fallah A, Goudarzi A, Pouladi I. Dengue virus: Etiology, epidemiology, pathobiology, and developments in diagnosis and control – A comprehensive review. Vol. 127, Infection, Genetics and Evolution. Elsevier B.V.; 2025.

2. Almeida MT de, Merighi DGS, Visnardi AB, Gonçalves CAB, Amorim VM de F, Ferrari AS de A, et al. Latin America’s Dengue Outbreak Poses a Global Health Threat. Viruses 2025, Vol 17, Page 57 [Internet]. 2025 Jan 1 [cited 2025 Feb 5];17(1):57. Available from: https://www.mdpi.com/1999-4915/17/1/57/htm

3. Alied M, Endo PT, Aquino VH, Vadduri VV, Huy NT. Latin America in the clutches of an old foe: Dengue. The Brazilian Journal of Infectious Diseases. 2023 Jul 1;27(4):102788.

4. Pan American Health Organization. PAHO/WHO Data - Dengue [Internet]. 2025 [cited 2025 Feb 8]. p. 1. Available from: https://www3.paho.org/data/index.php/en/mnu-topics/indicadores-dengue-en.html

5. Gallego-Munuera M, Colomé-Hidalgo M. Letalidad por dengue y desigualdades en la Región de las Américas entre el 2014 y el 2023. Revista Panamericana de Salud Pública [Internet]. 2024 Dec 18 [cited 2025 Feb 5];48:1. Available from: https://iris.paho.org/handle/10665.2/62938

6. Organización Mundial de la Salud. Dengue y dengue grave [Internet]. 20251 [cited 2025 Feb 13]. p. 1. Available from: https://www.who.int/es/news-room/fact-sheets/detail/dengue-and-severe-dengue

7. Tsheten T, Clements ACA, Gray DJ, Adhikary RK, Furuya-Kanamori L, Wangdi K. Clinical predictors of severe dengue: a systematic review and meta-analysis. Infect Dis Poverty [Internet]. 2021;10(1):123. Available from: 10.1186/s40249-021-00908-2

8. Chen HJ, Tang HJ, Lu CL, Chien CC. Warning signs and severe dengue in end-stage renal disease dialysis patients. Journal of Microbiology, Immunology and Infection [Internet]. 2020;53(6):979–85. Available from: https://www.sciencedirect.com/science/article/pii/S1684118219301379

9. Basto-Abreu A, López-Olmedo N, Rojas-Martínez R, Aguilar-Salinas CA, Moreno-Banda GL, Carnalla M, et al. Prevalencia de prediabetes y diabetes en México: Ensanut 2022. Salud Publica Mex [Internet]. 2023 Jun 13;65:s163–8. Available from: https://www.saludpublica.mx/index.php/spm/article/view/14832

10. Campos-Nonato I, Oviedo-Solís C, Hernández-Barrera L, Márquez-Murillo M, Gómez-Álvarez E, Alcocer-Díaz L, et al. Detección, atención y control de hipertensión arterial. Salud Publica Mex [Internet]. 2024 Aug 22;66(4, jul-ago):539–48. Available from: https://saludpublica.mx/index.php/spm/article/view/15867

11. Institute for Health Metrics and Evaluation (IHME). GBD Compare [Internet]. 2021 [cited 2025 Feb 13]. p. 1. Available from: https://vizhub.healthdata.org/gbd-compare/

12. Yang CH, Leeid IK, Chen YC, Huang WC, Hsu JC, Tai CH, et al. Prognostic factors in severe dengue patients: A multi-center retrospective cohort study. Samy AM, editor. PLoS Negl Trop Dis [Internet]. 2025 Jan 28 [cited 2025 Feb 5];19(1):e0012846. Available from: https://journals.plos.org/plosntds/article?id=10.1371/journal.pntd.0012846

13. Mehta P, Hotez PJ. NTD and NCD Co-morbidities: The Example of Dengue Fever. Vol. 10, PLoS Neglected Tropical Diseases. Public Library of Science; 2016.

14. Ríos-Bracamontes EF, Mendoza-Cano O, Lugo-Radillo A, Ortega-Ramírez AD, Murillo-Zamora E. Factors Contributing to In-Hospital Mortality in Dengue: Insights from National Surveillance Data in Mexico (2020–2024). Trop Med Infect Dis. 2024 Sep 3;9(9):202.

15. Instituto de Diagnóstico y Referencia Epidemiólogicos. Lineamientos para la Vigilancia por Laboratorio. Dengue y otras arbovirosis [Internet]. Ciudad de México; 2021 [cited 2025 Feb 13]. Available from: https://www.gob.mx/cms/uploads/attachment/file/629265/Lineamientos_Dengue_Arb_V1-2021.pdf

16. Wang JY, Tseng CC, Lee CS, Cheng KP. Clinical and upper gastroendoscopic features of patients with dengue virus infection. J Gastroenterol Hepatol [Internet]. 1990 Dec 1;5(6):664–8. Available from: 10.1111/j.1440-1746.1990.tb01122.x

17. Agência Brasil. Dengue cases in Brazil surpass 6.4M in 2024 [Internet]. 2025 [cited 2025 Feb 13]. p. 1–2. Available from: https://agenciabrasil.ebc.com.br/en/saude/noticia/2025-01/dengue-cases-brazil-surpass-64m-2024

18. Gobierno de México. Programa sectorial de salud 2020-2024 [Internet]. 2020 [cited 2025 Feb 17]. p. 1–42. Available from: https://dof.gob.mx/nota_detalle.php?codigo=5598474&fecha=17/08/2020#gsc.tab=0

19. Bambrick HJ, Capon AG, Barnett GB, Beaty RM, Burton AJ. Climate Change and Health in the Urban Environment: Adaptation Opportunities in Australian Cities. Asia Pacific Journal of Public Health [Internet]. 2011 Jan 17;17(2_suppl):67S–79S. Available from: 10.1177/1010539510391774

20. Lachlan M, Rokho K, Alistair W, Simon H, Jeffery S, Dianne K, et al. Health Impacts of Climate Change in Pacific Island Countries: A Regional Assessment of Vulnerabilities and Adaptation Priorities. Environ Health Perspect [Internet]. 2016 Nov 1;124(11):1707–14. Available from: 10.1289/ehp.1509756

